# Rare but elevated incidence of hematological malignancy after clozapine use in schizophrenia: a population cohort study

**DOI:** 10.1101/2024.08.07.24311592

**Authors:** Yuqi Hu, Le Gao, Lingyue Zhou, Wenlong Liu, Cuiling Wei, Boyan Liu, Qi Sun, Wenxin Tian, Rachel Yui Ki Chu, Song Song, Franco Wing Tak Cheng, Joe Kwun Nam Chan, Amy Pui Pui Ng, Heidi Ka Ying Lo, Krystal Chi Kei Lee, Wing Chung Chang, William Chi Wai Wong, Esther Wai Yin Chan, Ian Chi Kei Wong, Yi Chai, Francisco Tsz Tsun Lai

## Abstract

**Background:** Recent disproportionality analyses and nationwide case-control studies suggested a potential association between clozapine use and hematological malignancy (HM). Nevertheless, the absolute rate difference is unclear due to the absence of cohort studies.

**Methods:** We extracted data from a territory-wide public healthcare database in Hong Kong to build a retrospective cohort of anonymized patients aged 18+ with a diagnosis of schizophrenia who used clozapine or olanzapine (drug comparator with highly similar chemical structure and pharmacological mechanisms) for 90+ days, with at least two prior other antipsychotic use records within both groups. Weighted by inverse probability of treatment based on propensity scores, Poisson regression was used to estimate the incidence rate ratio (IRR) of HM between clozapine and olanzapine users. The absolute rate difference was also estimated.

**Results:** In total, 9,965 patients were included, with 834 clozapine users, who had a significant IRR of 2.22 (95% CI 1.52, 3.34) for HM compared to olanzapine users. Absolute rate difference was estimated to be 57.40 (95% CI 33.24, 81.55) per 100,000 person-years. Findings were consistent across sub-groups by age and sex in terms of effect size, although the IRR was non-significant for those aged 65 or older. Sensitivity analyses all supported the robustness of the results and showed good specificity to HM but no other cancers.

**Conclusion:** Absolute rate difference in HM incidence was very small although there is a twofold elevated rate. Pharmacotherapies with clozapine may consider this potential rare risk in addition to known side effects.

## INTRODUCTION

The rare yet elevated incidence of cancer associated with antipsychotic use, particularly breast cancer, has been noted in previous research.^1-3^ However, randomized controlled trials investigating these relatively rare long-term safety outcomes are oftentimes infeasible. Of note, real-world evidence on the risk of hematological malignancy (HM) associated with the use of clozapine, an atypical antipsychotic agent, is accruing. A disproportionality analysis of VigiBase® in 2020 initially provided preliminary analytical evidence on such an association for clozapine.^4^ This finding underscores the need for further pharmacovigilance actions.^5^ Previous research has shown that clozapine is associated with hematological abnormalities, such as agranulocytosis, eosinophilia and thrombocytopenia.^6^ In 2022, a nationwide Finnish study used a population-based case-control design and suggested a threefold increase in the odds of HM after using clozapine compared to other antipsychotics.^7^ Similarly, a recent case-control study using the United States Veteran Health Administration Database showed comparable results.^8^ Due to their retrospective ascertainment of antipsychotic use, however, the comparison between specific antipsychotic agents is not without limitations. Though there was also a cohort study design in the Finnish study, it was used to estimate the crude incidence rate of HM without multivariable analyses. The absolute rate difference, e.g., number of HM cases associated with clozapine use per 100,000 person years, remains to be estimated.

Moreover, clozapine is typically initiated at a different level of drug resistance in patients compared to most other antipsychotics.^9^ In fact, it is the only drug approved by the U.S. Food and Drug Administration for treatment-resistant schizophrenia.^10^ Hence, before using clozapine, patients have likely used other antipsychotics, and indication bias or confounding by other drugs is highly plausible. Unfortunately, the comparison between clozapine and other drugs on a comparable timeframe was not feasible in the retrospective case-control approach to the ascertainment of drug use. In light of this potential bias, an analytic cohort study that takes into consideration previous drug use as a proxy for the level of drug resistance is needed to substantiate the findings. Of equal importance, a cohort study can provide an estimate of the potential absolute rate difference to better inform clinical decisions.

In this study, we took advantage of a territory-wide public healthcare database in Hong Kong with comprehensive linkage to various healthcare attendance records, diagnoses and medication use among people with schizophrenia. We aimed to test for the association of clozapine use with HM in comparison with olanzapine users, adjusting for prior other antipsychotic use within both groups. Olanzapine was chosen as a comparator due to its similar chemical structure and increasing advocacy in recent years for it to serve as an alternative to clozapine for treatment-resistant schizophrenia.^11,12^ We hypothesize an elevated risk of HM following the use of clozapine compared with olanzapine.

## METHODS

### Data source

We adopted a retrospective cohort study design for this study. The Hospital Authority of Hong Kong, which is the sole provider of public inpatient services and a major provider of public outpatient services, provided strictly anonymized and de-identified electronic health records for data analysis. As all legal residents of Hong Kong are eligible for receiving services from the public sector, the database essentially covers the entire population and all territories of Hong Kong. The electronic health records were extracted from the Clinical Management System in which clinicians input routine patient attendance, diagnosis and medication records daily, linked by a unique person identity number. Diagnoses are coded based on the International Classification of Diseases, Ninth Revision, Clinical Modifications (ICD-9-CM), while medications are coded with British National Formulary codes as well as generic names, with dosage and duration information also available. Timestamps are available for every record entry, facilitating the delineation of the temporality of records and events. Many pharmacoepidemiologic research studies have already been published using this database,^13-16^ with the accuracy of diagnostic codes well established.^17^

### Participants

We included anonymized patients aged 18 years or older with a diagnosis of schizophrenia (ICD-9-CM code 295) and a record of using clozapine or olanzapine for a duration of 90 days or more in January 2001 -August 2022. The first prescription of clozapine or olanzapine date was designated as the index date. Patients who i.) did not use at least two other antipsychotics before the index date, ii.) those who ever used both clozapine and olanzapine in their prescription history, and iii.) those who had a prior record of any cancers(including HM) before the index date were excluded. Records dating back to the year 1999 were used to execute these exclusion criteria. Patients were followed from the index date (i.e., 90 days or more clozapine or olanzapine use) until i.) the diagnosis of HM, ii.) death, iii.) five years after the cessation of olanzapine or clozapine use, or iv.) end of data availability (i.e., August 31^st^ 2022), whichever came earliest.

Ethics approval for this study was obtained from the Institutional Review Board of the University of Hong Kong/ Hospital Authority Hong Kong West Cluster (HKU/HA HKW IRB, reference number: UW 20-113). Informed consent was waived as a requirement for ethics approval because the data were all anonymized.

### Outcomes

Any HM diagnosis (ICD-9-CM codes 200-209, 238.4, 238.5, 238.6, 238.7, detail in **eTable 1**) as a composite measure was adopted as the outcome of this study.

**eTable 1.** ICD-9-CM diagnostic codes and generic drug names used to define covariates.

| Disease name/ Drug category | ICD-9-CM codes/ drug names |
| --- | --- |
| Hematological malignancy |  |
| Lymphosarcoma and reticulosarcoma | 200 |
| Hodgkin's disease | 201 |
| Other malignant neoplasms of lymphoid and histiocytic tissue | 202 |
| Multiple myeloma and immunoproliferative neoplasms | 203 |
| Lymphoid leukaemia | 204 |
| Myeloid leukaemia | 205 |
| Monocytic leukaemia | 206 |
| Other specified leukaemia | 207 |
| Leukaemia of unspecified cell type | 208 |
| Polycythemia vera | 238.4 |
| Neoplasm of uncertain behavior of histiocytic and mast cells | 238.5 |
| Neoplasm of uncertain behavior of plasma cells | 238.6 |
| Neoplasm of uncertain behavior of other lymphatic and hematopoietic tissue | 238.7 |
| Mental illnesses |  |
| Depression | 296.2, 296.3, 300.4, 625.4, 293.83, 311, 648.4, 300.0 |
| Bipolar disorder | 296 |
| Dementia | 290, 294.1, 294.2, 294.8, 331.0, 331.1, 331.82 |
| Immunological disease | 42, 43, 44, 266.2, 273.0, 273.1, 273.2, 277.2, 277.6, 279, 288.1, 289.89 |
| Autoimmune Diseases: |  |
| Vitiligo | 709.01 |
| Addison disease | 255.4 |
| Alopecia areata | 704.01 |
| Autoimmune/Hashimoto's thyroiditis | 245.2 |
| Graves' disease | 242 |
| Morphoea | 701 |
| Multiple sclerosis | 340 |
| Myasthenia gravis | 358 |
| Pernicious anaemia | 281 |
| Primary biliary cirrhosis | 571.6 |
| Takayasu arteritis | 446.7 |
| Type 1 diabetes mellitus | 250.X1, 250.X3 |
| Associated Conditions: |  |
| Episcleritis/scleritis | 379.0X |
| Erythema nodosum | 695.2 |
| Haemolytic anaemia | 283.XX |
| Immune thrombocytopenia purpura | 287.31 |
| Leukocytoclastic vasculitis | 446.29 |
| Myositis | 729.1 |
| Pulmonary fibrosis/interstitial lung disease | 515. X, 516.31 |
| Raynaud's syndrome | 443 |
| Sjögren's syndrome/sicca syndrome | 710.2 |
| Thrombocytopenia purpura | 287.3 |
| Vasculitis | 447.6 |
| Juvenile idiopathic arthritis (JIA) | 714.30, 714.31, 714.32, 714.33, 714.2 |
| Acquired immune deficiencies | 42, 43, 44 |
| Asthma | 493 |
| Previous antipsychotic use |  |
| Phenothiazines | Chlorpromazine, Perphenazine, Fluphenazine, Trifluoperazine, Pericyazine, Thioridazine |
| Thioxanthene derivatives | Flupenthixol, Zuclopenthixol, Thiothixene |
| Diphenylbutylpiperidine derivatives | Pimozide |
| Butyrophenone derivatives | Haloperidol, Droperidol |
| Benzamides | Sulpiride, Amisulpride |
| Indole derivatives | Lurasidone, Ziprasidone, Molindone, Sertindole |
| Diazepines, oxazepines, thiazepines and oxepines | Quetiapine, Asenapine |
| Other antipsychotics | Aripiprazole, Brexpiprazole, Risperidone, Paliperidone |

### Exposure and comparator

Clozapine and olanzapine were identified by their generic names in the electronic health records. We did not consider a 7-day, or a shorter, gap between repeated prescriptions of antipsychotics as a discontinuation given a half-life of olanzapine ranging from 21 to 54 hours.^18^ Clozapine was the primary exposure in this study while olanzapine was the comparator.

### Statistical analysis

Poisson regression was used to estimate the incidence rate ratio (IRR) with 95% confidence intervals (CI) of HM between clozapine and olanzapine users, with an offset term to account for varying follow-up times for each patient. The inverse probability of treatment weighting (IPTW), using a propensity score, was applied to balance the characteristics between the clozapine and olanzapine users. The score was estimated by the logistics regression, considering covariates including age, sex, previous use of categories of antipsychotics according to their chemical structure (phenothiazines, thioxanthene derivatives, diphenylbutylpiperidine derivatives, butyrophenone derivatives, benzamides, indole derivatives, diazepines, oxazepines, thiazepines and oxepines, and other antipsychotics), prior mental disorder diagnoses (depression, bipolar disorder, and dementia), number of antipsychotic used in the past year, and immunological disease history (autoimmune disease and acquired immune deficiencies) which have been proven to associate with the HM also used as covariates^19,20^. **eTable 1** shows the ICD-9-CM codes and generic drug names used to ascertain the covariates. The standardized mean difference (SMD) was used to identify potential imbalances between clozapine and olanzapine users. Covariates with an SMD greater than 0.1 after IPTW were further adjusted in the regression model. In addition to IRRs, weighted absolute rate differences were also estimated between the two groups.

Subgroup analyses by age group and sex were conducted to provide specific estimates for different demographic strata. Additionally, several sensitivity analyses were conducted to test the robustness of our results: i) the cohort was further restricted to patients using clozapine or olanzapine for 180 days (instead of 90); ii) patients were censored for three years (instead of five) after their cessation of clozapine or olanzapine use; iii) a multivariable Poisson regression to employed to replicate the analysis; iv) a Cox regression with the IPTW approach was used to estimate the hazard ratio; and v) asthma (ICD-9-CM codes 493) and other cancers (ICD-9-CM codes 140-239, excluding HM) were used as negative control outcomes to check for any unmeasurable selection bias, as it shared the same potential source of bias with HM but was not causally related to antipsychotic exposure.

YH and LG were independent analysts of this study to minimize errors in coding and programming.

## RESULTS

We identified 444,716 people from the database who had a prescription of antipsychotics between January 2001 and August 2022. After applying the exclusion criteria, the cohort size was 9,965, comprising 834 clozapine users and 9,131 olanzapine users. **Figure 1** shows the flowchart of the cohort selection procedure.

**Figure 1.**
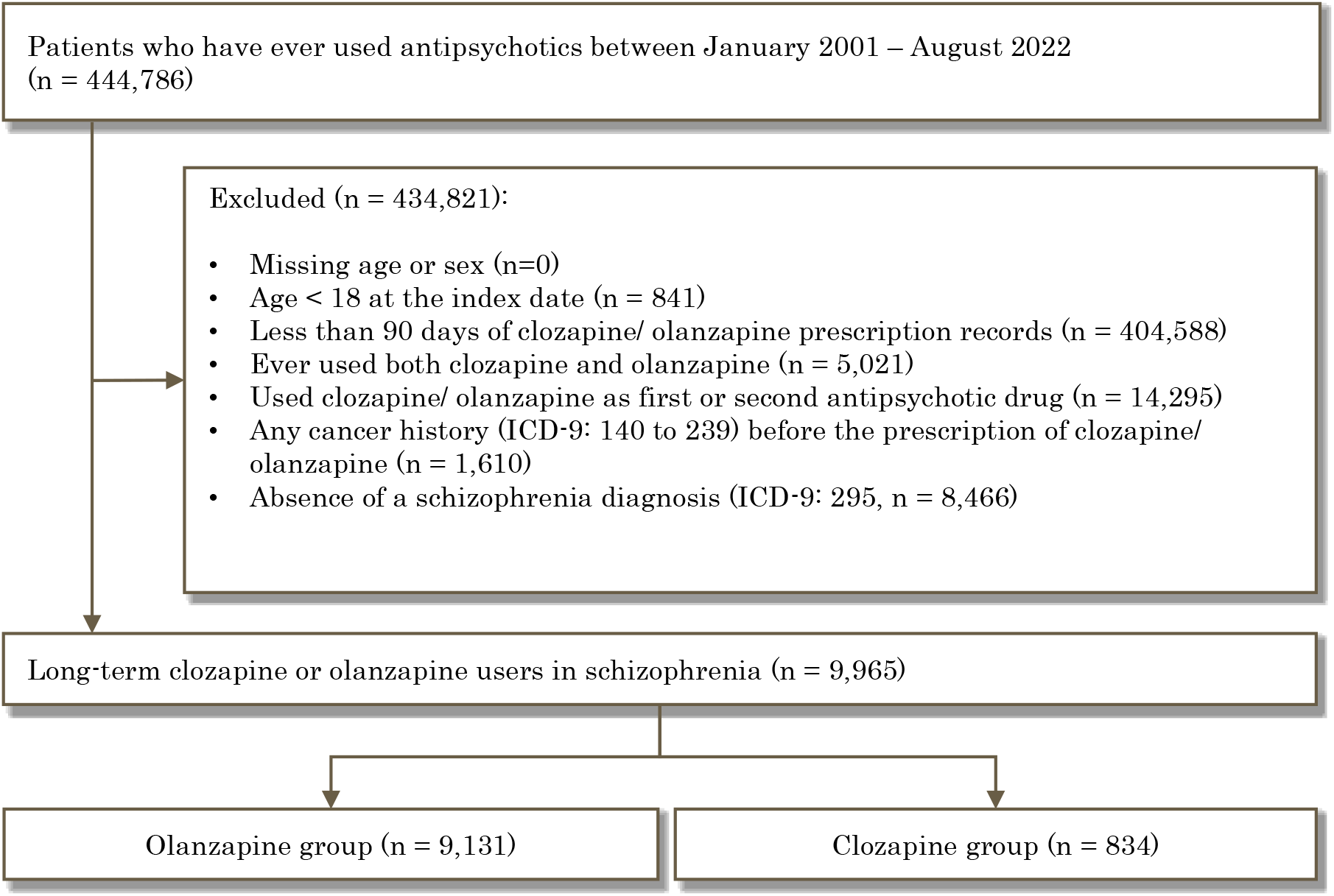
Flow chart showing cohort selection processes.

### Cohort characteristics

**Table 1** shows the baseline characteristics of the cohort before and after applying IPTW. Before weighting, the mean age of the clozapine group (41.00 ± 15.02) was approximately five years younger than that of the olanzapine group. The olanzapine group had a higher proportion of men (54.0%) than women, while clozapine the group had a smaller proportion of men (46.6%). On average, the follow-up time of the clozapine users were 4540.90 days compared to 2664.43 days follow-up time in olanzapine users, and more clozapine users prescribed more than three kinds of other antipsychotics in the past year (59.5% versus 36.9%). Notable differences were observed in the types of previous antipsychotics use. Specifically, phenothiazines (82.6% versus 64.6%) and thioxanthene derivatives (52.8% versus 37.4%) were more often used previously among clozapine users than olanzapine users. However, more olanzapine users had used diazepines, oxazepines, thiazepines, and oxepines (39.5% versus 28.4%), and ‘other antipsychotics’ (71.9% versus 57.3%). Additionally, a higher prevalence of psychiatric comorbidities and immunological diseases was observed among olanzapine users. After weighting, the SMD of characteristics were all less than 0.1 except prior benzamide use, which was adjusted in the subsequent weighted regression analysis.

**Table 1.** Cohort characteristics.

|  | Unweighted |  |  | Weighted by inverse probability of treatment |  |  |
| --- | --- | --- | --- | --- | --- | --- |
|  | Olanzapine | Clozapine | SMD <sup>a</sup> | Olanzapine | Clozapine | SMD <sup>a</sup> |
| *N* | 9131 | 834 |  | 9964.8 | 10101.1 |  |
| Mean age (standard deviation) | 46.48 (15.0) | 41 (11.5) | 0.404 | 46.0 (14.9) | 44.8 (12.9) | 0.084 |
| Sex (%) |  |  | 0.144 |  |  | 0.036 |
| Men | 4932 (54) | 389 (46.6) |  | 4643 (46.6) | 4541 (45) |  |
| Women | 4199 (46) | 445 (53.4) |  | 5321 (53.4) | 5560 (55) |  |
| Number of other antipsychotic drugs used in the past year |  |  | 0.515 |  |  | 0.024 |
| 0 | 215 (2.4) | 22(2.6) |  | 237 (2.4) | 245 (2.4) |  |
| 1 | 2203 (24.1) | 81(9.7) |  | 2284 (22.9) | 2229 (22.1) |  |
| 2 | 3348 (36.7) | 235(28.2) |  | 3583 (36.0) | 3724 (36.9) |  |
| ≥3 | 3365 (36.9) | 469(59.5) |  | 3861 (38.7) | 3903 (38.6) |  |
| Previous other antipsychotics use (%) |  |  |  |  |  |  |
| Phenothiazines | 5902 (64.6) | 689 (82.6) | 0.413 | 6591 (66.1) | 6493 (64.3) | 0.043 |
| Thioxanthene derivatives | 3413 (37.4) | 440 (52.8) | 0.316 | 3853 (38.7) | 3638 (36) | 0.057 |
| Diphenylbutylpiperidine derivatives | 222 (2.4) | 44 (5.3) | 0.150 | 265 (2.7) | 288 (2.8) | 0.011 |
| Butyrophenone derivatives | 6532 (71.5) | 621 (74.5) | 0.069 | 7155 (71.8) | 7245 (71.7) | 0.001 |
| Benzamides | 3949 (43.2) | 364 (43.6) | 0.013 | 4320 (43.4) | 4903 (48.5) | 0.103 |
| Indole derivatives | 536 (5.9) | 65 (7.8) | 0.076 | 602 (6.0) | 753 (7.5) | 0.057 |
| Diazepines, oxazepines, thiazepines and oxepines | 3605 (39.5) | 237 (28.4) | 0.232 | 3843 (38.6) | 4239 (42) | 0.071 |
| Other antipsychotics | 6563 (71.9) | 478 (57.3) | 0.312 | 7043 (70.7) | 7400 (73.3) | 0.059 |
| Mental illness diagnoses (%) |  |  |  |  |  |  |
| Depression | 1132 (12.4) | 61 (7.3) | 0.170 | 1194 (12.0) | 1283 (12.7) | 0.048 |
| Bipolar disorder | 1118 (12.2) | 73 (8.8) | 0.118 | 1192 (12.0) | 1362 (13.5) | 0.006 |
| Dementia | 154 (1.7) | 2 (0.2) | 0.148 | 156 (1.6) | 165 (1.6) | 0.050 |
| History of immunological diseases (%) | 248 (2.7) | 18 (2.2) | 0.032 | 265 (2.7) | 191 (1.9) | 0.028 |

<sup>a</sup> Standardized mean difference

### Main analysis

Figure 2. shows the weighted cumulative incidence curves of HM over the follow-up period for clozapine and olanzapine users. The incidence of HM among clozapine users was consistently higher than that among olanzapine users for nearly the entire follow-up period.

**Figure 2.**
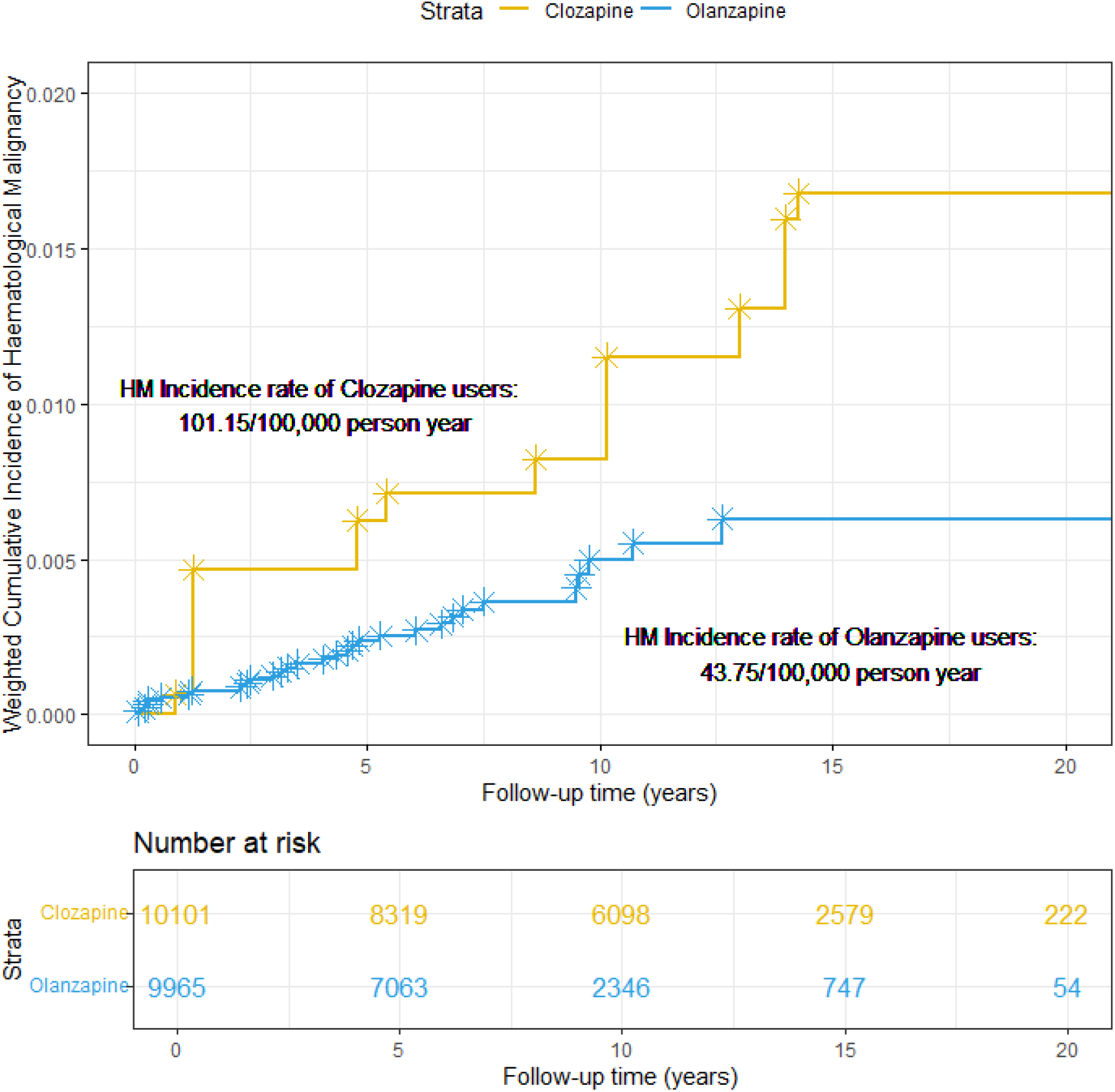
Cumulative incidence curves over the observation period by clozapine or olanzapine use status. Yellow represents clozapine and blue represents olanzapine.

Among all 9,965 individuals exposed to olanzapine or clozapine, a total of 39 individuals were diagnosed with HM. Out of the 39 cases, nine individuals were clozapine users (n = 834), while the remaining 30 individuals were olanzapine users (n = 9,131). Six clozapine users and 23 in olanzapine users developed HM after discontinuing the medication. For the six clozapine users, the average number of days between their discontinuation and HM was 256.67 days. For the 23 olanzapine users, the number was 713.86 days.

Results from the Poisson regression models show that compared to olanzapine users, clozapine users had a significantly higher IRR of 2.22 (95% CI 1.52, 3.34) for HM (**Table 2**). The absolute rate difference of HM between the two groups was 57.40 (95% CI 33.24, 81.55) per 100,000 person-years.

**Table 2.** Incidence rate ratios (IRR) of hematological malignancy (HM) with 95% confidence intervals (CI)

|  | Cohort size | Average follow-up duration (days) | Number of HM cases | Crude incidence rate of HM per 100,000 person-year | Weighted incidence rate of HM per 100,000 person-year | Crude IRR (CI) | Weighted IRR (CI) | Weighted absolute rate difference per 100,000 person-years (CI) |
| --- | --- | --- | --- | --- | --- | --- | --- | --- |
| Main analysis | 9,965 | 2821.47 | 39 | 50.66 | 78.01 |  |  | 57.40 (33.24,81.55) |
| Olanzapine | 9,131 | 2664.43 | 30 | 45.04 | 43.75 | Ref. | Ref. |  |
| Clozapine | 834 | 4540.90 | 9 | 86.80 | 101.15 | 1.93 (0.86,3.90) | 2.22 (1.52,3.34) |  |
| Men | 4,644 | 2763.26 | 18 | 51.23 | 76.01 |  |  | 46.04 (9.57,82.52) |
| Olanzapine | 4,199 | 2581.79 | 15 | 50.54 | 48.98 | Ref. | Ref. |  |
| Clozapine | 445 | 4475.57 | 3 | 55.02 | 95.02 | 1.09 (0.25,3.30) | 2.33 (1.33,4.31) |  |
| Women | 5,321 | 2872.28 | 21 | 50.19 | 72.26 |  |  | 54.27 (23.41,85.14) |
| Olanzapine | 4,932 | 2734.78 | 15 | 40.62 | 39.34 | Ref. | Ref. |  |
| Clozapine | 389 | 4615.64 | 6 | 122.06 | 93.61 | 3.00 (1.07,7.39) | 2.35 (1.38,4.24) |  |
| Aged 44 or younger | 4,821 | 3196.96 | 8 | 18.96 | 27.93 |  |  | 23.49 (4.33,42.64) |
| Olanzapine | 4,301 | 2980.84 | 5 | 14.24 | 13.75 | Ref. | Ref. |  |
| Clozapine | 520 | 4984.57 | 3 | 42.27 | 37.24 | 2.97 (0.61,12.10) | 2.71 (1.15,7.67) |  |
| Aged 45 - 64 | 3,967 | 2620.41 | 22 | 77.30 | 110.06 |  |  | 72.69 (25.06,120.31) |
| Olanzapine | 3,674 | 2518.13 | 17 | 67.12 | 66.96 | Ref. | Ref. |  |
| Clozapine | 293 | 3902.97 | 5 | 159.7 | 139.65 | 2.38 (0.78,6.01) | 2.07 (1.24,3.65) |  |
| Aged 65 or older | 1,177 | 1961.14 | 9 | 142.41 | 192.19 |  |  | 137.22 (0.00,301.95) |
| Olanzapine | 1,156 | 1952.16 | 8 | 129.48 | 129.34 | Ref. | Ref. |  |
| Clozapine | 21 | 2455.48 | 1 | 708.33 | 266.55 | 5.47 (0.29,29.81) | 1.90 (0.62,5.78) |  |

Subgroup analyses showed a significantly elevated risk of HM among clozapine users for all sex and age groups, except for those aged 65 or older despite a comparable IRR.

### Sensitivity analysis

Sensitivity analyses yielded similar results (**eTable 2)**. The IRR for asthma as a negative control outcome was not significant and close to one (IRR 0.90, 95% CI 0.66,1.21). Likewise, the IRR for other cancers was estimated at 0.77 (95% CI 0.68, 0.86).

**eTable 2.** Sensitivity analysis: incidence rate ratios (IRR) of hematological malignancy (HM) with 95% confidence intervals (CI)

|  | Cohort size | Average follow-up duration (days) | Number of HM cases | Crude incidence rate of HM per 100,000 person-year | Weighted incidence rate of HM per 100,000 person-year | Crude IRR (HR for Cox regression) | Weighted IRR (HR for Cox regression) | Weighted absolute rate difference per 100,000 person-years (CI) |
| --- | --- | --- | --- | --- | --- | --- | --- | --- |
| Restricting to those who used clozapine or olanzapine for 180+ days | 6,091 | 2781.40 | 21 | 45.27 | 59.46 |  |  | 34.79 (6.94,62.65) |
| Olanzapine | 5,529 | 2618.33 | 16 | 40.37 | 38.85 | Ref. | Ref. |  |
| Clozapine | 562 | 4385.72 | 5 | 74.09 | 73.65 | 1.84 (0.6,4.69) | 2.29 (1.29, 4.07) |  |
| Right-censoring patients at 3 years after treatment discontinuation | 9,965 | 2709.44 | 39 | 52.76 | 79.99 |  |  | 56.53 (31.76,81.29) |
| Olanzapine | 9,131 | 2546.58 | 30 | 47.12 | 45.79 | Ref. | Ref. |  |
| Clozapine | 834 | 4492.47 | 9 | 87.74 | 102.32 | 1.93 (0.86,3.91) | 2.22 (1.32, 4.19) |  |
| Multivariable Poisson regression | 9,965 | 2821.47 | 39 | 50.66 | 78.01 |  |  | - |
| Olanzapine | 9,131 | 2664.43 | 30 | 45.04 | 43.75 | Ref. | Ref. |  |
| Clozapine | 834 | 4540.90 | 9 | 86.80 | 101.15 | 1.93 (0.86,3.90) | 2.53 (1.04, 5.42) |  |
| Weighted Cox regression | 9,965 | 2821.47 | 39 | 50.66 | 78.01 |  |  | 57.40 (33.24,81.55) |
| Olanzapine | 9,131 | 2664.43 | 30 | 45.04 | 43.75 | Ref. | Ref. |  |
| Clozapine | 834 | 4540.90 | 9 | 86.80 | 101.15 | 1.95 (0.91, 4.19) | 2.28 (0.80, 6.48) |  |
| Using asthma as negative control outcome | 9,814 | 2817.21 | 74 | 97.76 | 106.17 |  |  | 8.73 (0.00, 37.93) |
| Olanzapine | 8,989 | 2660.68 | 64 | 97.74 | 100.30 | Ref. | Ref. |  |
| Clozapine | 825 | 4522.73 | 10 | 97.89 | 110.20 | 1.01 (0.52, 1.97) | 0.90 (0.66,1.21) |  |
| Using other cancers as negative control outcome | 9,982 | 2747.85 | 597 | 794.98 | 644.99 |  |  | 175.53 (97.68, 253.3) |
| Olanzapine | 9,140 | 2595.88 | 497 | 765.1 | 749.89 | Ref. | Ref. |  |
| Clozapine | 842 | 4397.43 | 70 | 690.52 | 574.35 | 0.90 (0.70, 1.15) | 0.77 (0.68,0.86) |  |

## DISCUSSION

Findings from this territory-wide cohort study involving approximately 10,000 individuals with schizophrenia in Hong Kong suggested a rare but twofold increased rate of HM associated with clozapine compared to olanzapine. Nevertheless, the absolute rate difference is small, with less than 60 additional cases of HM per 100,000 person years of clozapine use compared with olanzapine use. Potential confounding factors, such as previous other antipsychotic use, underlying mental illnesses, and history of immunological diseases observed at the baseline, were properly adjusted. Subgroup analyses by age and sex, as well as sensitivity analyses, showed highly robust results, supporting a comparably strong and highly specific association. Our research hypothesis is therefore well supported by the data.

### Relationship with the literature

To the best of our knowledge, this work is the world’s first analytic cohort study on this association to estimate absolute rate differences, with only two prior notable case-control studies conducted in Finland^7^ and the United States.^8^ Although the Finnish study also provided descriptive data on the incidence of HM using a cohort design, no hypothesis testing was conducted in the cohort study. Indeed, our descriptive absolute risk estimates were highly comparable with the Finnish study’s descriptive cohort.^7^ The rarity of HM and the relatively low prevalence of clozapine use compared to some other widely used antipsychotics have both greatly limited previous research on this association, despite compelling evidence on the association between clozapine use and a range of hematological abnormalities.^21-23^ In fact, apart from this and the two case-control studies, existing evidence on an association with HM arose only from passive reporting systems using disproportionality analysis which may be subject to more biases than large electronic health record epidemiologic studies due to known limitations in adverse event reporting platforms and databases.^5,24^ The previous case-control studies, despite the strengths of the national registry and comprehensive veteran health database, used retrospectively ascertained long-term clozapine use as the main exposure and ‘little to no such use’ as the comparator. This approach is not without limitations, as clozapine use is most typically preceded by a range of other antipsychotic use in the individuals because it is widely regarded as the most efficacious drug available.^25^ Our current study address this challenge by confining our cohort to those who used two other antipsychotics before using clozapine (or olanzapine) and employing olanzapine as an active comparator.^26^ This ensures a more proper adjustment of previous antipsychotic use before clozapine or olanzapine initiation. With a cohort design, we also estimated the absolute rate difference which is highly relevant in clinical decision making. The current study represented evidence that is one level higher up the clinical evidence pyramid compared with the existing knowledge, i.e., from case-control to cohort evidence.

### Interpretation of observed association

The strong association observed in this and the previous two case-control studies^7^ as well as its good coherence with previous evidence on the hematological abnormalities^27^ associated with clozapine constitute a strong case that this association is likely causal. We believe the effects of such abnormalities being induced by the prolonged use of clozapine may accumulate over the duration of use and potentially increase the risk of malignancy through a range of potential mechanisms.^28^ For instance, recent evidence showed that the elevated risk of agranulocytosis following clozapine use may persist well over several years, and this mechanism could potentially impact the increased risk of other blood-related disorders among clozapine users, including a higher risk of HM.^29^ We did not have a sufficient number of cases to separately analyze the type of HM with a more significantly elevated risk; inference about the specific underlying mechanism is thus limited. Further research should investigate specific HM types.

### Potential clinical implications

Despite a small rate difference, a potential strong association between clozapine use and HM, if substantiated by further evidence, would provide additional information to the safety profile of the drug compared to olanzapine. This potential slight recalibration of the risk-benefit balance may partially impact clinical decisions about long-term clozapine use. First, considering the markedly elevated risk of HM, despite its rare incidence, psychiatrists may be more inclined to try other antipsychotics before initiating clozapine, which is currently the most efficacious antipsychotic drug. Second, for individuals with significant hematological abnormalities detected early in the course of clozapine use, alternative drugs may be beneficial, as prolonged use of clozapine may be associated with a higher chance of HM. Third, patients with a family history of HM may benefit from reconsideration of clozapine prescription, as they have a higher baseline risk.^30^ These potential considerations need to be supported by additional evidence, such as studies specific to individuals with a family history of HM and the relationship between detected hematological abnormalities and subsequent HM risks.

### Strengths and limitations

One of the key strengths of this study is the cohort design with a range of important covariates properly adjusted, e.g., prior antipsychotic use, providing highly useful information on the absolute rate of HM and the rate difference. Additionally, the territory-wide database with comprehensive medical records based on the same coding system and practices also confers great strengths, such as good generalizability and representativeness of the findings.

There are, nevertheless, limitations that warrant caution. First, this evidence is observational without randomization, which may entail potential selection or indication bias. Nevertheless, blood tests prior to the prescription of clozapine should have already excluded people with a higher risk of HM; such indication bias, if any, might lead to an underestimation of the observed association. Moreover, it is difficult to conduct randomized prospective studies on such a rare outcome, so future research should likely still be focused on observational studies with multiple sites or even countries. Second, although olanzapine, particularly in high doses, can be useful for some cases, it is not a suggested replacement option for clozapine in the treatment-resistance schizophrenia due to the lack of a highly comparable indication.^31^ Third, there are unobserved covariates such as socioeconomic status and lifestyle factors that were not included in this study. Fourth, there may be a detection bias of HM for clozapine, as it is known to be associated with hematological abnormalities which are closely monitored.^32^ However, since we allowed the observation to last until five years after discontinuation to account for symptomatic presentation, the detection bias should not be significant. Also, the association is strong and unlikely to be purely arising from detection bias, i.e., earlier detection. In fact, our findings showed that a smaller proportion of HM cases was recorded during the use of clozapine than that during the use of olanzapine. Fifth, the dosage of treatment was not standardized during the course of treatment or across individuals and is subject to the psychiatrists’ clinical judgement for each individual. Sixth, the follow-up time for the olanzapine group was shorter than that of the clozapine group, but it should likely be long enough to observe and capture the study outcome, with a mean follow-up time of more than seven years. Seventh, among clozapine users, many had used olanzapine before switching to clozapine in clinical practice but we excluded approximately 5,000 patients who had a history of using olanzapine either before or after starting clozapine treatment. This exclusion has limited our sample size although we arrived at a more selective sample for a fair comparison. Finally, the study population in Hong Kong is predominantly ethnic Chinese, and the generalizability of the findings to other populations need to be tested using multinational data.

## CONCLUSIONS

To conclude, we conducted a territory-wide retrospective cohort study in Hong Kong and identified a rare but evidently elevated risk of HM, i.e., twofold, among clozapine users with schizophrenia compared with olanzapine users. If substantiated by further studies, the risk-benefit ratio of the long-term prescription of clozapine should be slightly recalibrated.

## Data Availability

The data underlying the results presented in the study are available from Hong Kong Hospital Authority's Central Panel on Administrative Assessment of External Data Requests (https://www3.ha.org.hk/data/Provision/Submission).

## CONTRIBUTORS’ STATEMENT

Yuqi Hu, Le Gao, Yi Chai and Francisco Lai contributed to the conception of the work. All authors designed the study. Yuqi Hu and Le Gao contributed to the acquisition and analysis of the data, and all authors interpreted the data. Yuqi Hu and Francisco Lai drafted the manuscript. All authors revised the manuscript critically for important intellectual content, gave final approval of the version to be published and agreed to be accountable for all aspects of the work. Yuqi Hu and Le Gao are co-first authors. Yi Chai and Francisco Lai share the senior authorship, had full access to all the data in the study and took final responsibility for the decision to submit for publication.

## ACKNOWLEDGEMENTS

The authors thank the Hospital Authority for the generous provision of data and gratefully acknowledge Professor Martin Roland of the University of Cambridge for his invaluable advice.

